# HIV status is not associated with SARS-CoV-2 viral load and longer duration of infection among people with well-controlled HIV and high COVID-19 vaccine coverage

**DOI:** 10.1101/2025.05.11.25327398

**Authors:** Didhiti Deb, Mohammad Eghbal Heidari, Balladiah Kizito, Monamodi Kesamang, Tebogo Matila, Maitshwarelo Matsheka, Stefan Niemann, Sanghyuk S. Shin, Chawangwa Modongo

## Abstract

**BACKGROUND:** HIV may alter SARS-CoV-2 viral load and increase the duration of SARS-CoV-2 infection and, thereby, drive the emergence of variants of concern. This study investigated the association between HIV with SARS-CoV-2 viral load and duration of infection in Botswana, in the era of high antiretroviral therapy (ART) coverage.

**METHODS:** This cohort study was conducted in Gaborone, Botswana, covering 6 healthcare facilities. Gaborone residents with confirmed SARS-CoV-2 infection were recruited through active screening, passive diagnosis, and contact tracing. Among enrolled participants, we conducted weekly serial testing for SARS-CoV2. The primary outcomes for this analysis were SARS-CoV-2 viral load and duration of SARS-CoV-2 infection. SARS-CoV-2 viral load was measured using RT-PCR cycle threshold (Ct) value at the time of diagnosis with higher Ct value indicating a lower viral load. Duration of SARS-CoV-2 infection was calculated as the number of days between symptom onset and date of negative PCR test. Multiple linear regression and Cox-proportional hazard models were employed to determine the association between HIV infection status and outcome variables.

**FINDINGS:** Of the 578 enrolled participants, 486 (84.1%) with known HIV status were included in the analysis. Of the population, 69.3% were female, mean age was 37.8 years, and 88.7% were vaccinated for COVID-19. People living with HIV (PLWH) accounted for 26.1% of the population, with a mean CD4+ T cell count of 715 cells/µL. Among PLWH, 86.6% were taking ARTs at the time of diagnosis. The mean Ct value across all participants was 22.5 (SD: 6.16). No significant difference in Ct values was observed between HIV and CD4+ T cell categories. A significant association was found between age and Ct values (β = −0.05, 95% CI: −0.11, 0.00, p = 0.03). Survival analysis showed increased time to negative COVID-19 PCR for PLWH with CD4+ T cell count ≥ 500 cells/µL (HR = 1.45, 95% CI: 1.02, 2.05, p = 0.03) compared to people without HIV, but not for PLWH CD4+ T cell count < 500 cells/µL (HR = 1.01, 95% CI: 0.62, 1.65, p = 0.9).

**INTERPRETATION:** Among PLWH with well-controlled HIV disease and high vaccine coverage, we found no evidence of an association between HIV status with SARS-CoV-2 viral load or increased duration of SARS-CoV-2 infection. Additional research is needed to confirm shorter duration of SARS-CoV-2 infection among PLWH with CD4+ T cell counts ≥ 500 cells/µL.

**SOURCE OF FUNDING:** This research was supported by US NIH grant #R01AI170204.

## Introduction

The COVID-19 pandemic, caused by the severe acute respiratory syndrome coronavirus 2 (SARS-CoV-2), has posed significant challenges to global health systems, economies, and societies(1). Although the virus indiscriminately affected populations worldwide, specific subgroups, particularly those with underlying chronic health conditions, may experience differential outcomes. Among these, individuals living with human immunodeficiency virus (HIV) in regions with a high prevalence of HIV/AIDS represent a critical group for investigation. In southern Africa, the dual epidemics of HIV and COVID-19 have created a compounded public health challenge(2). The potential influence of HIV on SARS-CoV-2 transmission dynamics and its evolution within the host remains a critical area of exploration(3,4).

HIV-associated immunosuppression could alter the host’s ability to suppress SARS-CoV-2 replication and, thus, affect viral load concentrations within the host. SARS-CoV-2 viral load is an important predictor of infectiousness and onward transmission. HIV infection could also affect the ability of the host to clear SARS-CoV-2 infection, resulting in persistent infection and increased opportunities for onward transmission(5–7). Persistent SARS-CoV-2 infection may also lead to the emergence of variants of concern. Notably, in the case of tuberculosis, HIV status remained a predictor of *Mycobacterium tuberculosis* bacterial load even when antiretroviral therapy (ART) was widely available(8).

Understanding how HIV affects the duration of COVID-19 illness is critical for public health. Prolonged symptom duration or viral shedding in people living with HIV (PLWH) may increase their risk of transmitting SARS-CoV-2 to others, particularly in regions with high population density or limited healthcare access. Additionally, extended SARS-CoV2 replication in immunocompromised individuals could foster conditions conducive to SARS-Cov2 variants evolution, potentially leading to the emergence of new variants of concern(9). In this study, we examine the associations between HIV status with SARS-CoV-2 viral load and duration of infection, which could offer critical insights into COVID-19 transmission dynamics in populations with a high prevalence of HIV.

## Methods

### Study Design, Setting, and Participants

We analyzed data from a prospective cohort study to determine the association between HIV status with SARS-CoV-2 viral load and infection duration, respectively. Data were collected as part of an ongoing genomic epidemiology study conducted in Gaborone, the capital of Botswana, covering 13 healthcare facilities. Gaborone residents with confirmed SARS-CoV-2 infection without hearing, speaking, or cognitive difficulties were included. The study employed passive case-finding based on clinical presentations of SARS-CoV-2 symptoms, active case-finding encompassing universal patient screening, and contact tracing of diagnosed patients. Enrolled participants made weekly study visits with nasal samples taken for SARS-CoV-2 PCR testing at each visit until a negative test result.

Clinical and demographic data were collected through a standardized questionnaire, capturing information about SARS-CoV-2 symptoms and their duration. The difference between the date of sample collection and the date of first symptom onset is approximated as the time from symptoms to diagnosis. The time from symptoms to the last positive test is explained by the difference between the first PCR test based on the date of sample collection and the last positive PCR test. For survival analysis, time to event (time to negativity PCR) is determined as the difference between the first PCR test based on the date of sample collection and the last date that subjects were tested. Medical records were accessed to obtain data on SARS-CoV-2 and HIV history, including CD4 cell counts and antiretroviral therapy (ART) history. SARS-CoV-2 PCR results were recorded, including the cycle threshold (Ct) value for NGene and ORF1ab targets.

## Statistical Analysis

Standard descriptive statistics were performed, presenting frequencies for categorical variables and central tendencies for continuous variables. The Wilcoxon rank-sum test was used to compare Ct values across various HIV and CD4 categories. Spearman correlation was utilized to explore the relationship between CD4 cell counts and Ct values due to the non-normal distribution of the data.

To assess the association between HIV status and SARS-CoV-2 viral load in the context of Botswana, bivariate and multivariable linear regression models were employed. The outcome variable was the Ct value for SARS-CoV-2 PCR. The primary exposure was HIV status, categorized as HIV negative, HIV positive with CD4+ T cell count ≥ 500 cells/µl, and HIV positive with CD4+ T cell count < 500 cells/µl. Confounding variables included age, gender, smoking status, vaccination status, underlying diseases (except HIV), and history of COVID-19, which were derived from the existing literature review, prior knowledge, and drawing the directed acyclic graph (DAG).

For all multivariable models, all potential confounders were included regardless of their p-value in the model. We employed Kaplan-Meier analysis to estimate the survival functions and Cox regression to evaluate the impact of HIV status and other covariates on the time to negative PCR, defining the duration of SARS-CoV-2 infection from the time of diagnosis to the date of the last positive test. All analyses were performed using R statistical software version 4.3.1.

## Ethics Approval

The study received approval from the institutional review board at the University of California, Irvine and the Botswana Ministry of Health and Wellness Human Research Development Committee. Written consent was obtained from all participants.

## Results

### Study Demographics

Between July 2022 and July 2024, 578 participants were enrolled in the study. We excluded individuals with unknown HIV status from the analysis (n = 92; 15.9%). Of the included participants (n = 486), the median follow-up period was 15 days (IQR = 24); 215 had a negative SARS-Cov-2 test at the end of follow-up and 231 were censored with no confirmed negative date. Of the study population, 69.3% were female, and the mean age was 37.8 years (standard division [SD]: 14), and the median of 36.0 years with a range of 7 to 88 years (Table 1). Most participants were symptomatic (90.1%). PLWH accounted for 127 (26.1%) participants, with a mean CD4+ T cell count of 715 cells/µL (SD: 292 cells/µL); 18 (14.2%) and 3 (2.4%) PLWH participants had CD4+ T cell count of < 500 cells/µL and < 200 cells/µL, respectively. Among PLWH, 86.6% of participants were taking ART at the time of diagnosis. The mean time from symptoms to diagnosis was 3.5 days (SD: 4.2 days). Additionally, 92.1% of PLWH and 87.5% of participants without HIV had been vaccinated for COVID-19 (Table 1).

**Table 1:**
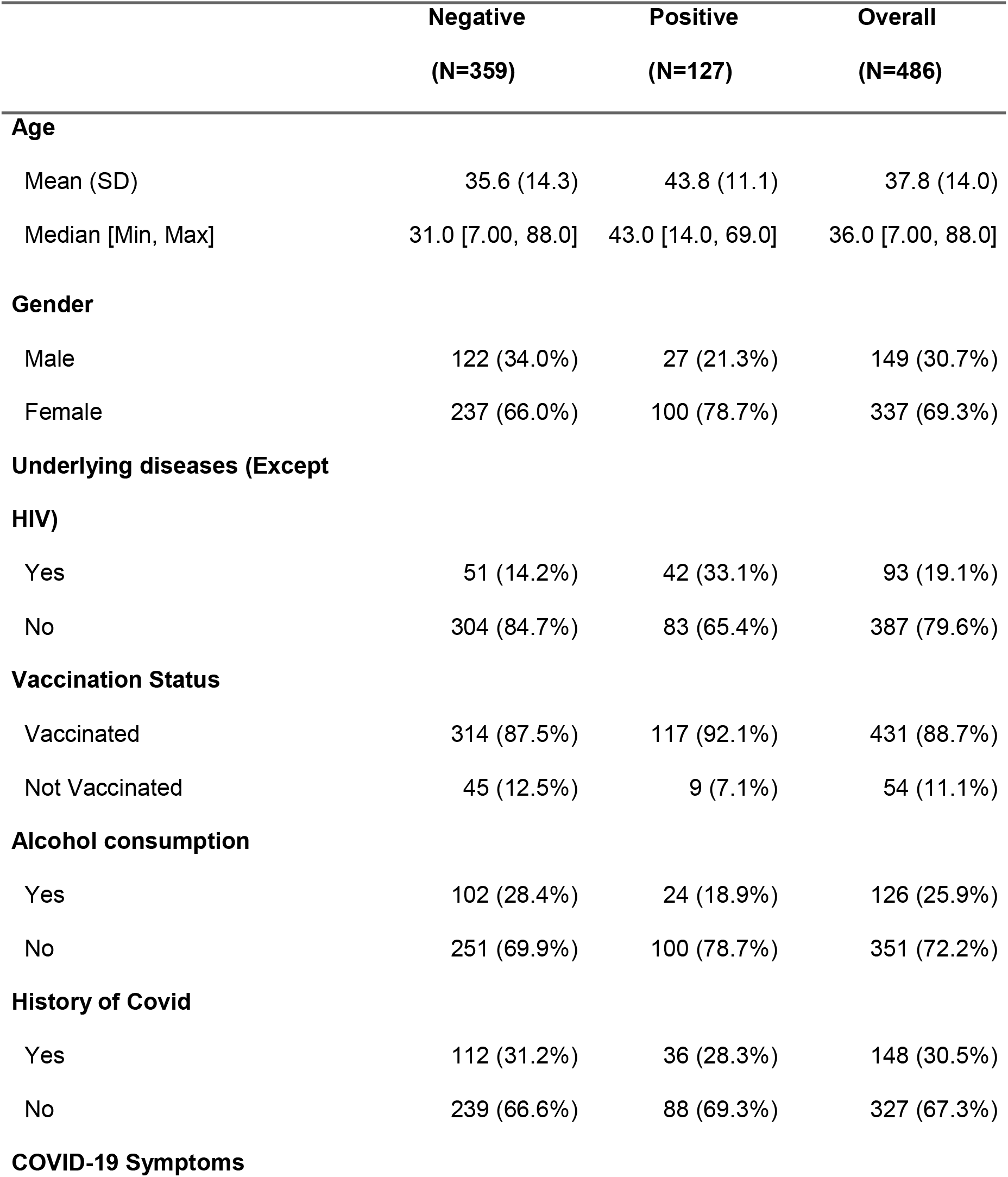
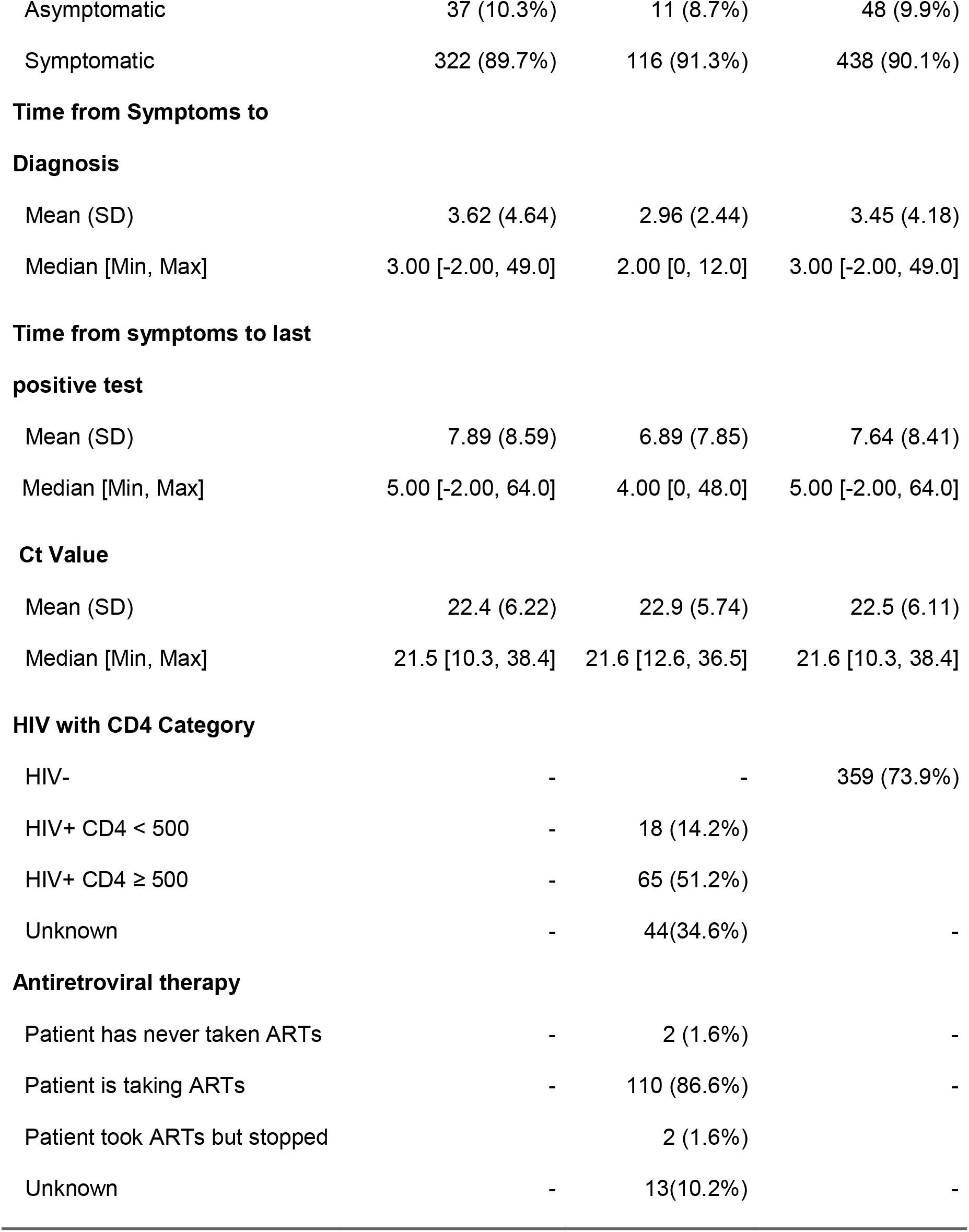
Demographic and Clinical Information by HIV Status of Individuals.

|  | **Negative**<br>**(N=359)** | **Positive**<br>**(N=127)** | **Overall**<br>**(N=486)** |
| --- | --- | --- | --- |
| **Age** |  |  |  |
| Mean (SD) | 35.6 (14.3) | 43.8 (11.1) | 37.8 (14.0) |
| Median [Min, Max] | 31.0 [7.00, 88.0] | 43.0 [14.0, 69.0] | 36.0 [7.00, 88.0] |
| **Gender** |  |  |  |
| Male | 122 (34.0%) | 27 (21.3%) | 149 (30.7%) |
| Female | 237 (66.0%) | 100 (78.7%) | 337 (69.3%) |
| **Underlying diseases (Except HIV)** |  |  |  |
| Yes | 51 (14.2%) | 42 (33.1%) | 93 (19.1%) |
| No | 304 (84.7%) | 83 (65.4%) | 387 (79.6%) |
| **Vaccination Status** |  |  |  |
| Vaccinated | 314 (87.5%) | 117 (92.1%) | 431 (88.7%) |
| Not Vaccinated | 45 (12.5%) | 9 (7.1%) | 54 (11.1%) |
| **Alcohol consumption** |  |  |  |
| Yes | 102 (28.4%) | 24 (18.9%) | 126 (25.9%) |
| No | 251 (69.9%) | 100 (78.7%) | 351 (72.2%) |
| **History of Covid** |  |  |  |
| Yes | 112 (31.2%) | 36 (28.3%) | 148 (30.5%) |
| No | 239 (66.6%) | 88 (69.3%) | 327 (67.3%) |
| **COVID-19 Symptoms** |  |  |  |
| Asymptomatic | 37 (10.3%) | 11 (8.7%) | 48 (9.9%) |
| Symptomatic | 322 (89.7%) | 116 (91.3%) | 438 (90.1%) |
| **Time from Symptoms to** |  |  |  |
| **Diagnosis** |  |  |  |
| Mean (SD) | 3.62 (4.64) | 2.96 (2.44) | 3.45 (4.18) |
| Median [Min, Max] | 3.00 [-2.00, 49.0] | 2.00 [0, 12.0] | 3.00 [-2.00, 49.0] |
| **Time from symptoms to last** |  |  |  |
| **positive test** |  |  |  |
| Mean (SD) | 7.89 (8.59) | 6.89 (7.85) | 7.64 (8.41) |
| Median [Min, Max] | 5.00 [-2.00, 64.0] | 4.00 [0, 48.0] | 5.00 [-2.00, 64.0] |
| **Ct Value** |  |  |  |
| Mean (SD) | 22.4 (6.22) | 22.9 (5.74) | 22.5 (6.11) |
| Median [Min, Max] | 21.5 [10.3, 38.4] | 21.6 [12.6, 36.5] | 21.6 [10.3, 38.4] |
| **HIV with CD4 Category** |  |  |  |
| HIV- | - | - | 359 (73.9%) |
| HIV+ CD4 < 500 | - | 18 (14.2%) |  |
| HIV+ CD4 ≥ 500 | - | 65 (51.2%) |  |
| Unknown | - | 44(34.6%) | - |
| **Antiretroviral therapy** |  |  |  |
| Patient has never taken ARTs | - | 2 (1.6%) | - |
| Patient is taking ARTs | - | 110 (86.6%) | - |
| Patient took ARTs but stopped |  | 2 (1.6%) |  |
| Unknown | - | 13(10.2%) | - |

### Association between HIV status and SARS-CoV-2 viral load

The mean Ct value was 22.5 (SD: 6.1) across all participants, and the median was21.6 ranging from 10.3 to 38.4 (Table 1). No significant difference in Ct values was observed between PLHW categorized by CD4 count and those without HIV (p = 0.80) (Figure 1).

**Figure 1.**
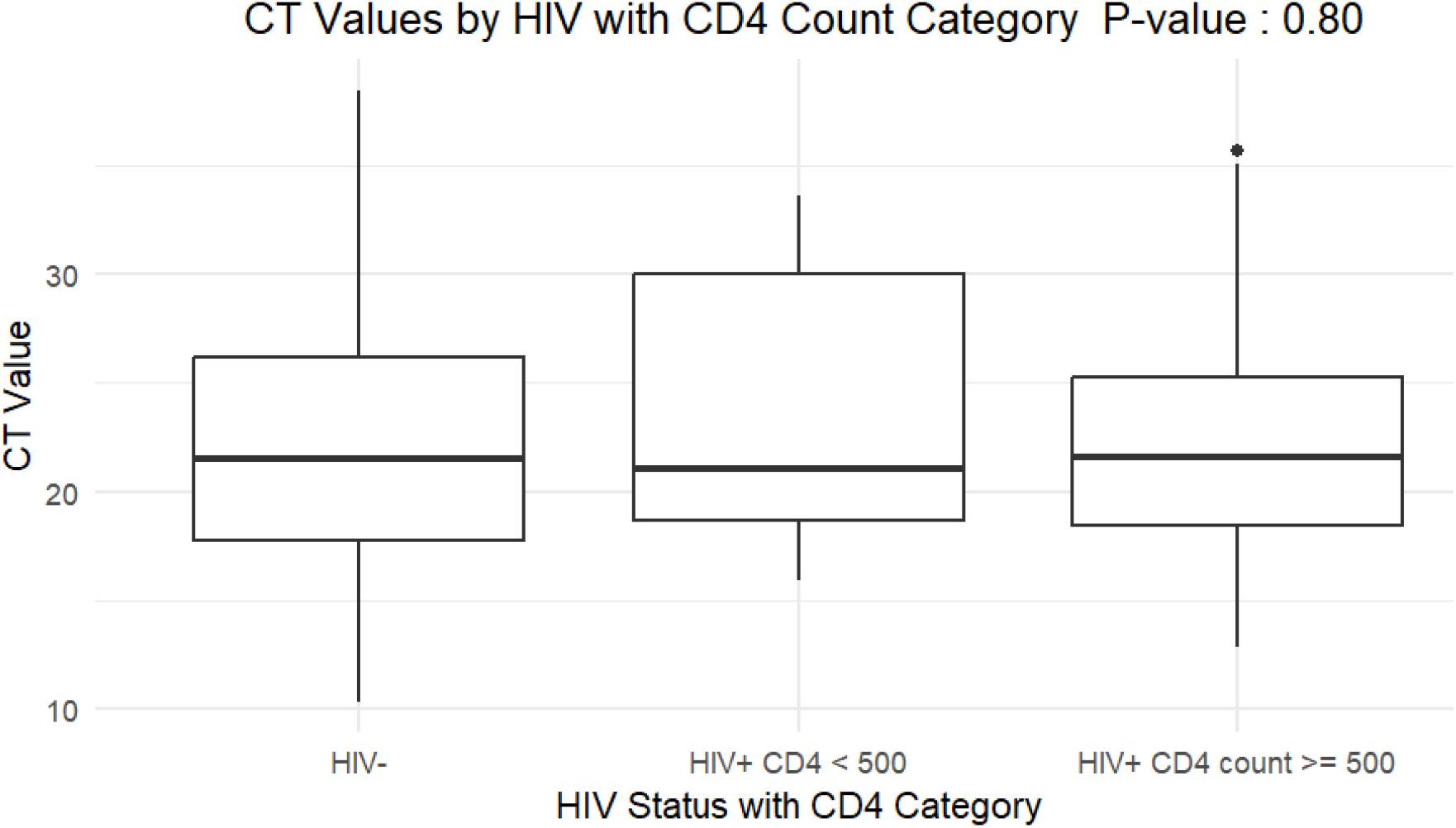
Boxplot showing the comparison of CT values across HIV-negative individuals and HIV-positive individuals by CD4 count categories (<500, 500–999, and ≥1000 cells/μL). The median CT values are similar across groups (P = 0.91).

**Figure 2.**
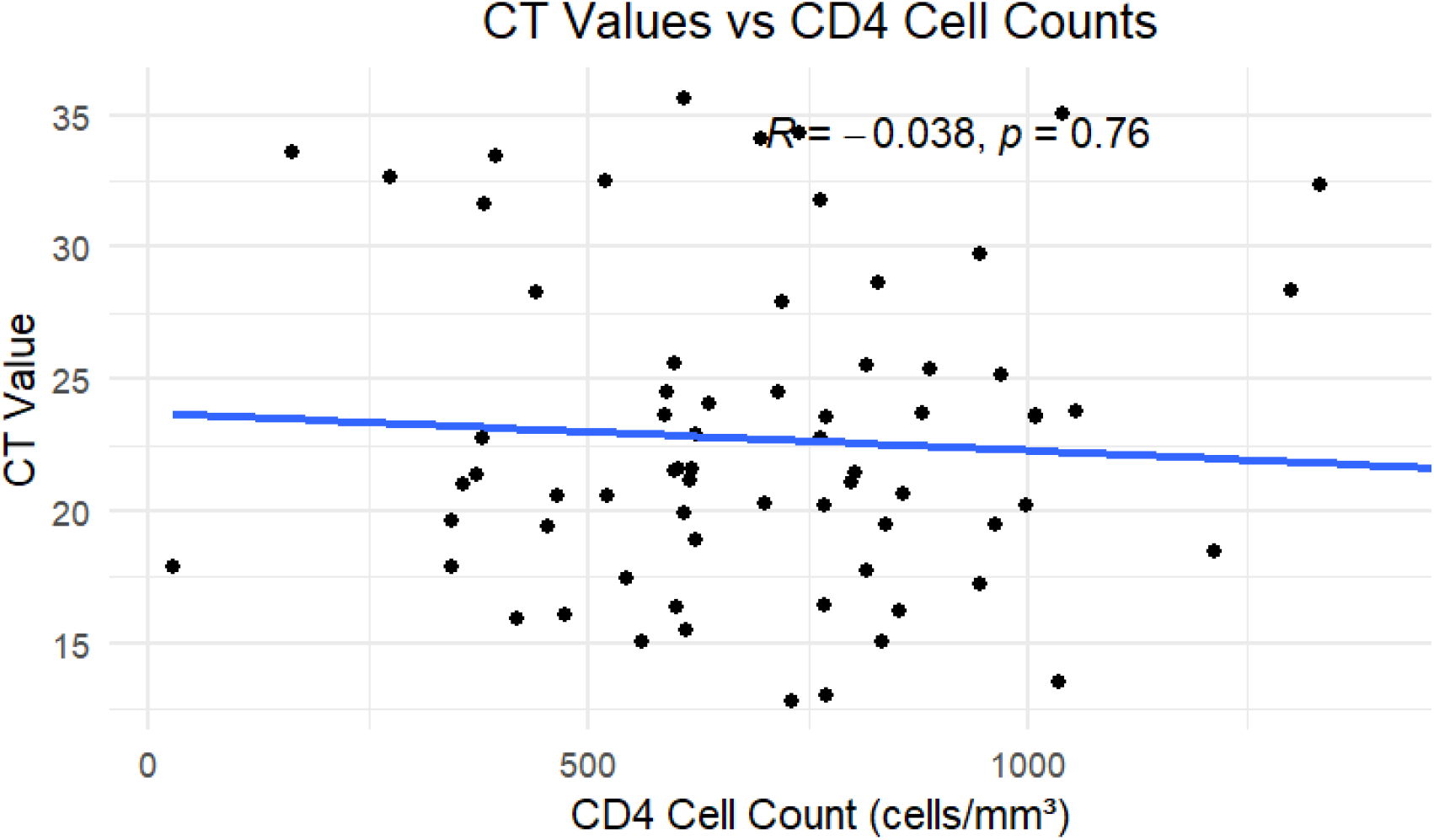
Scatter plot showing the relationship between CT values and CD4 cell counts (cells/µL).The blue line represents the linear regression line, with a correlation coefficient (R) of −0.038 and a p-value of 0.76.

**Figure 3.**
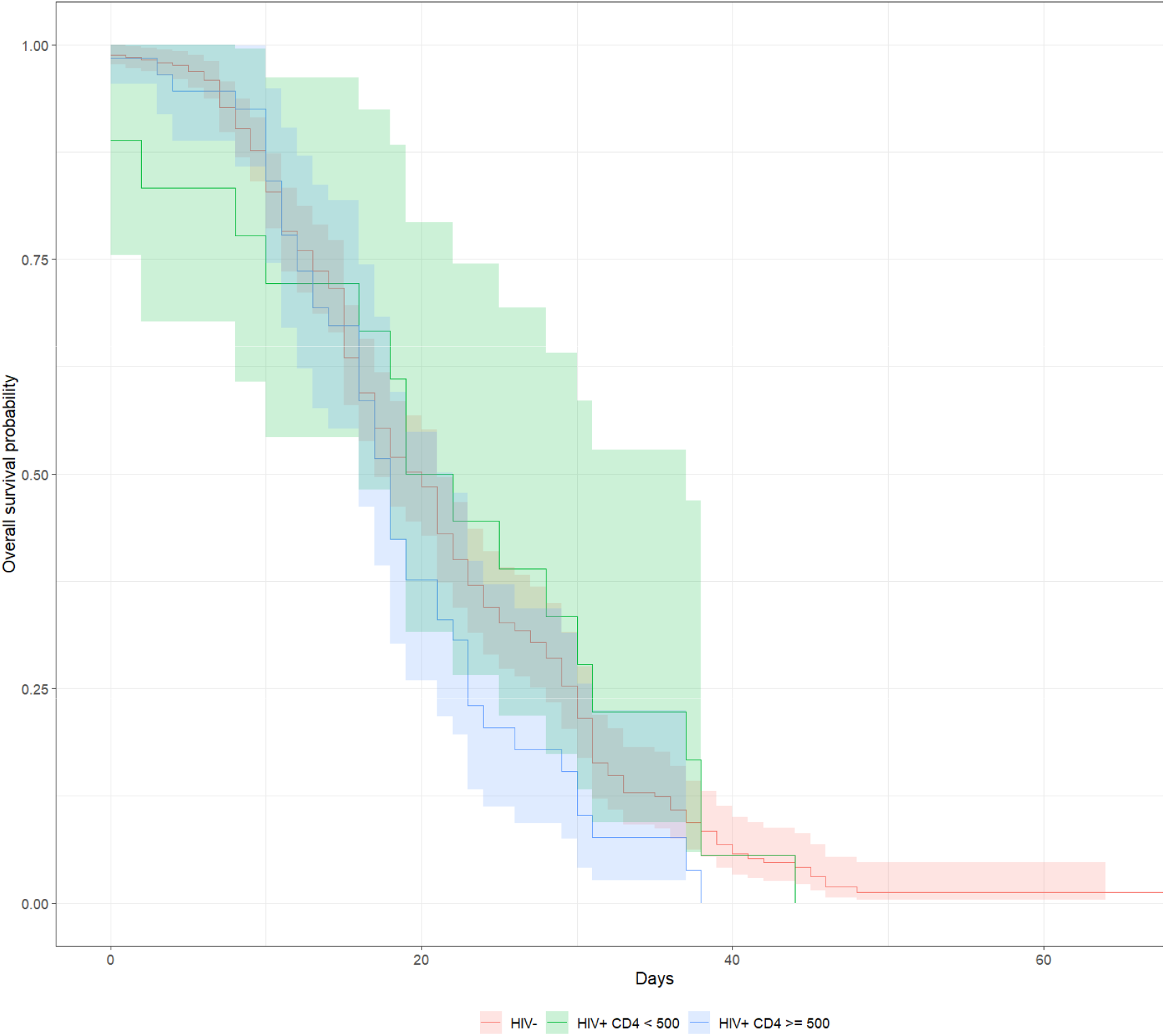
Kaplan-Meier Survival Curves comparing time-to-event outcomes between HIV with CD4 category.

Table 2 presents the results of the multivariable linear regression model assessing the association between HIV status and SARS-CoV-2 Ct values among participants. We found no evidence of an association between PLWH participants with CD4+ T cell counts < 500 cells/µL (beta = 1.4; 95% confidence interval [CI]: −1.90, 4.60; p-value = 0.4) or for participants with CD4 counts of 500 or higher (beta = 0.49; 95% CI: −1.40, 2.30; p-value = 0.6) compared to people without HIV. Age was significantly associated with Ct values, exhibiting a decrease with increasing age (beta = −0.05; 95% CI: −0.11, 0.00; p-value = 0.03).

**Table 2.**
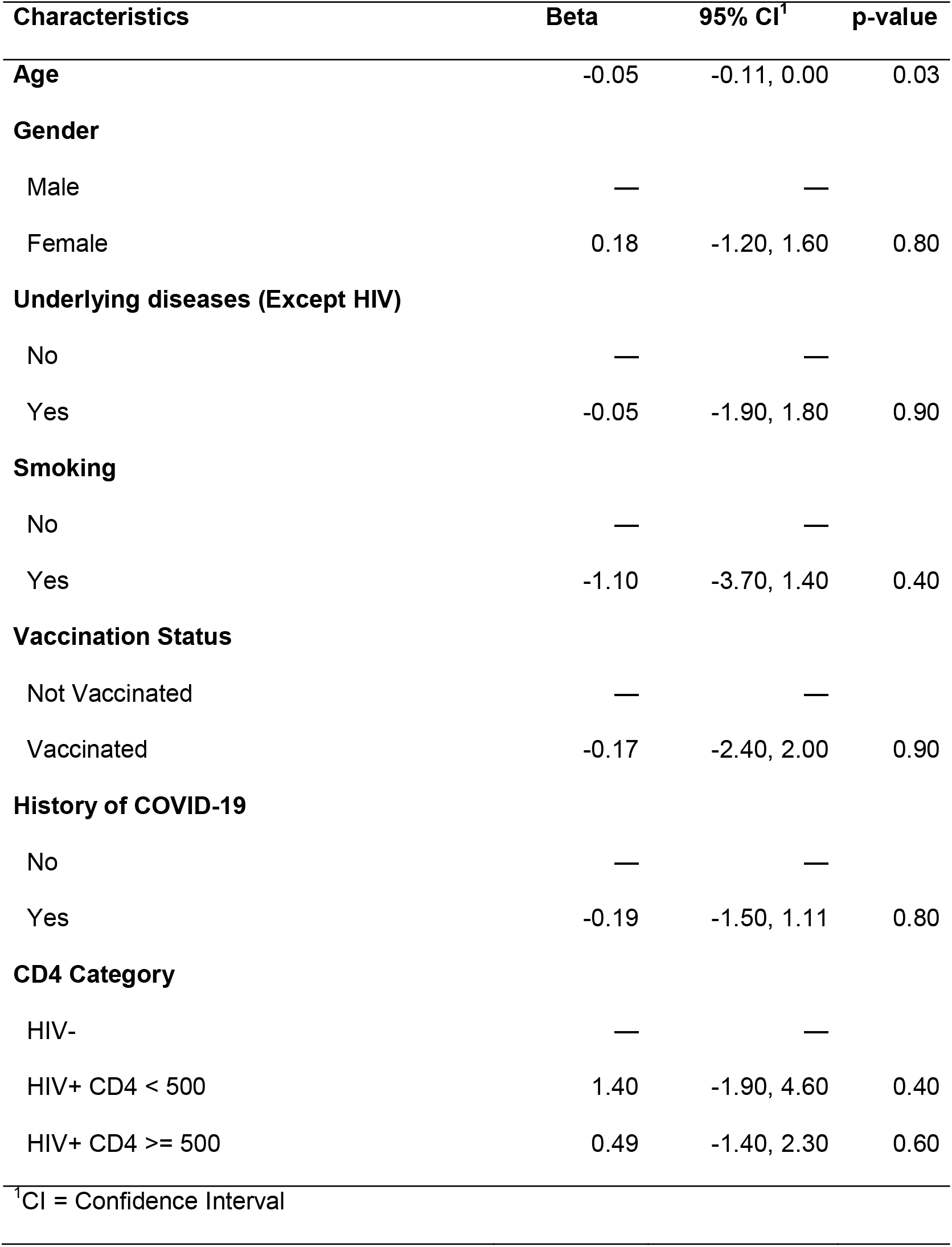
Multilinear Regression Model for Ct Value among all Participants.

### Survival analysis

Table 3 shows the results of the survival analysis, which evaluated the association between HIV status and time to negative SARS-CoV-2 PCR, while controlling for demographic and clinical confounders. The time to negative PCR for PLWH with CD4+ T cell count < 500 cells/µL was similar to that among people without HIV (hazard ratio [HR] = 1.01; p-value = 0.90). On average, PLWH with a CD4+ T cell count ≥ 500 cells/µL had shorter time to negative PCR compared to people without HIV (HR = 1.45; 95% CI 1.02, 2.05; p-value = 0.03).

**Table 3.** Survival Analysis of time to negativity.

| **Characteristics** | **HR<sup>1</sup>** | **95% CI<sup>1</sup>** | **p-value** |
| --- | --- | --- | --- |
| **Age** | 1.00 | 0.99, 1.01 | 0.70 |
| **Gender** |  |  |  |
| Female | — | — |  |
| Male | 0.95 | 0.73, 1.24 | 0.70 |
| **Smoker** |  |  |  |
| No | — | — |  |
| Yes | 0.80 | 0.50, 1.29 | 0.40 |
| **Underlying diseases (Except HIV)** |  |  |  |
| No | — | — |  |
| Yes | 0.81 | 0.58, 1.14 | 0.20 |
| **Vaccination Status** |  |  |  |
| Not Vaccinated | — | — |  |
| Vaccinated | 0.83 | 0.57, 1.21 | 0.30 |
| **History of COVID-19** |  |  |  |
| No | — | — |  |
| Yes | 1.17 | 0.91, 1.52 | 0.20 |
| **HIV with CD4 Category** |  |  |  |
| HIV- | — | — |  |
| HIV+ CD4 < 500 | 1.01 | 0.62, 1.65 | >0.9 |
| HIV+ CD4 ≥ 500 | 1.45 | 1.02, 2.05 | 0.03 |
| <sup>1</sup>HR = Hazard Ratio, CI = Confidence Interval |  |  |  |

## Discussion

In our study of COVID-19 in a setting with high vaccine coverage and well-controlled HIV we found no evidence of an association between HIV status or CD4+ T cell count with SARS CoV-2 viral load. Similarly, the median duration of SARS-CoV-2 shedding was 5 days and did not differ significantly by HIV status. However, survival analysis revealed that PLWH with CD4 counts > 500 cells/µL cleared the virus significantly faster than HIV-negative individuals.

Our findings of lack of association between HIV status and SARS-CoV-2 viral load are consistent with other studies that show that well-managed HIV patients may reduce the effect of HIV on SARS-CoV-2 viral load(10,11). Prior research has shown that PLWH on effective ART regimens might not experience more severe COVID-19 compared to the general population provided that their HIV is well controlled(12,13). Moreover, a systematic review highlighted that COVID-19 does not significantly impact the immunologic status or plasma HIV viral load in patients with HIV who are on stable ART(14). In contrast, a study on HIV with SARS-CoV-2 infection found that untreated HIV patients with extremely low CD4 counts may progress to severe outcomes when combined with SARS-CoV-2 infection(15). Because of their immune response history or the potential anti-retroviral treatment (ART) effectiveness against SARS-CoV 2, PLWH may even be protected from the most severe COVID-19 consequences.

Our findings are dissimilar with those from studies of tuberculosis, which have consistently found that HIV infection reduces *M. tuberculosis* bacterial load(8). One possible explanation for this difference is that the immune mechanisms controlling SARS-CoV-2 may be less dependent on CD4 T-cell function than those controlling tuberculosis. Acute SARS-CoV-2 infection is largely controlled by innate immune responses and B-cell/antibody-mediated immunity (along with CD8 T cells), especially in the early stages (16,17). In PLWH who have high CD4 counts and are virologically suppressed on ART, these arms of the immune system (neutralizing antibody production, innate antiviral responses) are mostly intact. Thus, their ability to control SARS-CoV-2 replication might remain comparable to HIV-negative individuals. Tuberculosis, on the other hand requires robust T-cell immunity (particularly Th1 CD4+ T-cells activating macrophages) for controlling bacterial growth. Conversely, decrease in CD4+ T-cell function can lead to reduced bacterial loads and altered disease presentation in HIV/TB co-infection, as *M. tuberculosis* exploits hyper-inflammatory host response to generate pathology in the lungs conducive to bacterial penetration in the airways. Another possibility is that a threshold effect might exist for HIV-associated immunosuppression in COVID-19: only when CD4 counts fall below a certain critical level (for example, < 200 or < 100 cells/µL) might HIV infection predispose to altered SARS-CoV-2 loads or prolonged viral shedding. In our cohort, nearly all PLWH had CD4 counts well above those severe immunosuppression levels, which could explain why we observed no difference in SARS-CoV-2 viral load by HIV status.

Case studies and genomic surveillance suggest that immunocompromised people, including PLWH, may carry SARS-CoV-2 infections for extended periods, encouraging within-host viral evolution and the creation of new variants of concern(5,16). In our study, no association was found between HIV status and duration of SARS-CoV-2 shedding. These findings are partly consistent with prior research from South Africa conducted prior to widespread access to COVID-19 vaccinations. For example, Meiring et al. (2022) reported no difference in overall shedding duration by HIV status but found that PLWH with CD4 <200 or unsuppressed HIV viral loads shed high viral load SARS-CoV-2 (Ct <30) significantly longer, with a median duration of 27 days compared to 7 days in HIV-negative individuals(17). Similarly, the PHIRST-C cohort study (Cohen et al., 2022) found that high SARS-CoV-2 viral load at baseline, immunosuppression, and symptomatic illness were associated with prolonged SARS-CoV-2 shedding(18). Kleynhans et al. (2023) also observed that while general HIV status did not affect episode length, immunosuppressed PLWH had longer high viral load shedding episodes(19). Coverage with full COVID-19 vaccination ranged from 0% to 25% in those studies. Our study extends those findings to a study population comprising mostly vaccinated individuals, including 92% vaccination coverage among PLWH.

Notably, our study found that PLWH with CD4+ T cell counts > 500 cells/µL demonstrated significantly faster viral clearance than HIV-negative individuals. This finding was not reported in prior studies of HIV-associated immunosuppression and time to viral clearance. Additional research is needed to confirm this finding and determine mechanisms for this association. Overall, these findings demonstrate that robust HIV care programs that achieve high coverage of ART may be important not just for individual-level outcomes, but also for public health initiatives to reduce SARS-CoV-2 transmission and evolution.

Our analysis also revealed that age was significantly associated with SARS-CoV-2 Ct values, with Ct values decreasing as age increased. Older age is consistently identified as a factor associated with higher viral loads in both the general population and among PLWH. This may be due to immune senescence, a decline in immune efficiency with age that impairs both innate and adaptive immune responses, resulting in slower viral clearance(20). Several studies, including Liu et al. (2020), have reported lower Ct values in patients over 50, indicating higher initial viral loads and worse clinical outcomes(21). Huang et al. (2020) and Nikolich-Žugich et al. (2020) further support that older adults experience more severe COVID-19, potentially due to elevated viral replication and prolonged shedding(22,23).

The study’s strengths lie in its rigorous methodological approach and the context of its setting—a region with a high HIV prevalence. This provides valuable epidemiological insights into the interplay between HIV and SARS-CoV-2. However, the study limitations are that the limited sample size of people with low CD4 counts (<200 cells/µL) restricts our ability to draw conclusions about the impact of advanced HIV-induced immunosuppression on SARS-CoV-2 viral load and duration of viral shedding.

Despite the limited number of participants with advanced immunosuppression (CD4 <200 cells/µL) in our cohort, our findings offer important insights into the relationship between immune status and SARS-CoV-2 viral dynamics in PLWH. The implication is that strong HIV care programs, marked by high ART coverage and successful immune reconstitution, can reduce both individual disease burden and broader public health risks, including potential SARS-CoV-2 transmission and within-host viral evolution. This challenges the notion that PLWH inherently pose a greater risk of sustaining high viral loads or contributing to the emergence of SARS-CoV-2 variants. Instead, our data suggest that well-managed HIV infection, with preserved CD4 counts and viral suppression, may limit such risks.

Previous studies have shown that severely immunocompromised individuals may carry chronic SARS-CoV-2 infections that facilitate within-host evolution of the virus, but have not adequately highlighted economic and other social and structural determinants of health—such as weak health systems, poverty, and stigma—that cause some PLWH to not be retained in care and thus to develop advanced immunodeficiency. Our findings suggest that such risks are a reflection of inadequate access to ART, weak health systems, and social inequities(20). Thus, strengthening HIV care systems not only benefits the health of PLWH but also represents a key strategy in pandemic preparedness and control. Promoting universal ART access and retention in care may serve as a dual intervention—mitigating HIV progression and reducing prolonged viral shedding of emerging pathogens like SARS-CoV-2.

## Data Availability

All data produced will be deposited to a public repository upon submission to journal

## Ethical Approval statement

The study received approval from the institutional review board at the University of California, Irvine, and the Botswana Ministry of Health and Wellness Human Research Development Committee. Written consent was obtained from all participants.

